# Healthcare professionals and point-of-care innovation: changing views and emerging trends

**DOI:** 10.1101/2025.05.03.25326929

**Authors:** Trevor Vigeant, Reilly Riddell, Bernard Ofosuhene, Grace White, Matheus Montenario, Ziyue Wang, Taylor Orwig, Heaven Y. Tatere, Bryan Buchholz, Denise Dunlap, David D. McManus, Ayorkor Gaba, Nathaniel Hafer

## Abstract

Point-of-care technologies (POCTs) have grown increasingly prevalent in clinical and at-home settings, offering various rapid diagnostic capabilities. This study presents findings from a nationwide survey of healthcare providers, including physicians, nurses, and allied health professionals, conducted between November 2023 and January 2024, capturing clinician perceptions of POCTs. This survey builds upon previous surveys from 2019 to 2021, and the data reflects a shift in attitudes toward a more neutral standpoint with fewer perceived benefits and concerns.

A total of 159 responses were analyzed. Core priorities, including accuracy, ease of use, and availability, remain consistently valued over the years. However, several perceived benefits, including continuous patient monitoring, diagnostic certainty, and patient management exhibited significant declines in agreement compared to previous years. This decline may be attributed to the increasing real-world experience with POCTs and the associated concerns surrounding inconsistent or inaccurate patient use, particularly in at-home settings. POCTs are now used for broader applications. Despite this, clinician perceptions of POCTs’ abilities to enhance patient-provider communication remained stable. The results suggest a tempered outlook among providers.

Evolving concerns may reflect heightened expectations and greater scrutiny as these technologies become commonplace. Agreement that POCTs may undermine clinical expertise increases, while concerns related to reimbursement and usability declines. Pilot questions related to artificial intelligence and machine learning were introduced; results indicated moderate openness to adopting AI-enhanced POCTs, particularly with tools offering novel clinical insights.

While the results provide valuable insight into an evolving landscape of clinician perceptions, limitations include a smaller sample size than in prior years and missing demographic data from a subset of participants. Future surveys will further integrate AI/ML-related questions while prioritizing broader demographic and geographic reach.

These trends underscore the importance of aligning the needs and expectations of stakeholders to enhance the clinical value of POCTs and improve patient outcomes.

## Introduction

Point-of-care technology (POCT) consists of tests conducted in clinical or at home environments for a range of applications, including blood glucose monitoring, home pregnancy testing, and over-the-counter COVID-19 testing [1]. POCTs have significantly evolved in recent years, playing an increasingly vital role in modern healthcare by enabling rapid diagnostics and enhancing patient management. Adopting these technologies has transformed clinical workflows by reducing turnaround times for results, facilitating immediate decision-making, and improving accessibility to critical diagnostic tools [2].

The COVID-19 pandemic served as a catalyst for widespread POCT implementation, emphasizing its importance in mitigating healthcare burdens, particularly in remote and resource-limited settings [3–5]. Between October 2021 and June 2022, 10.7 million self-administered COVID-19 tests were voluntarily reported by users of four manufacturers’ products. In the same period, 361.9 million laboratory-based and point-of-care test results were reported [6]. In the years following the pandemic, the focus has shifted from urgent deployment to refining and optimizing POCT integration within healthcare systems. Advancements aim to establish POCT as an indispensable component of modern healthcare, particularly in preventive medicine and chronic disease management [6]. Moreover, POCTs have expanded beyond infectious diseases to manage other noninfectious and chronic conditions including kidney disease, cancer, diabetes, and certain cardiovascular conditions [2, 3, 6].

This manuscript describes the results of a clinician-facing survey designed to measure clinician attitudes towards POCT use in clinical settings, adoption patterns, and perceived benefits and limitations over time. The survey was conducted from November 2023 to January 2024. Our analysis builds on previous surveys conducted between 2019 and 2021, which consistently highlighted three priorities of POCTs among healthcare professionals: accuracy, ease of use, and availability [1, 7]. Despite technological advancements, these priorities have remained unchanged, underscoring their continued relevance in guiding POCT innovation and adoption. By leveraging survey data collected over multiple years, we aim to assess trends in clinician attitudes, identify persistent barriers to implementation, and provide actionable insights for improving POCT integration. This survey aims to further investigate ongoing concerns while exploring new dimensions of POCT integration, including pilot questions pertaining to perspectives on artificial intelligence (AI) and machine learning (ML) applications.

## Materials and Methods

### Survey Development and Distribution

The 2023 POCT survey was developed following methodologies established in previous POCT surveys conducted in 2019, 2020 and 2021 [1, 3, 8]. It was developed to assess healthcare professionals’ perceptions of POCTs, including their importance, benefits, concerns, and clinical integration as their use-cases expand beyond traditional applications. The survey was distributed nationally to diverse healthcare professionals and researchers, ensuring a broad representation of clinical perspectives.

The distribution list included both internal and external email directories, academic medical centers (AMCs), and professional organizations. Contacts were obtained from large directories including the University of Massachusetts Center for Clinical Translational Science (UMCCTS), Massachusetts Medical Device Development Center (M2D2), UMass Memorial Health (UMMH), Consortia for Improving Medicine with Innovation and Technology (CIMIT), Center for Advancing Point of Care Technologies (CAPCaT), National Center for Complementary and Integrative Health (NCCIH), and other directories within the National Heart, Lung, and Blood Institute including the NIH Center for Accelerated Innovations (NCAI), Small Business Research Initiative (SBIR), and Research Evaluation and Commercialization Health (REACH) lists.

The survey was launched on 16 November 2023 and closed on 31 January 2024. It was distributed through email and via a LinkedIn invitation post. The exact number of individuals who received the email is unknown; however, based on the listserv populations, it is estimated to be over 15,000. To encourage response, reminder emails were sent monthly for the duration of the survey’s availability. To compensate respondents for their time, a $25 gift card was offered. The sole exclusion criterion was self-identification as a non-healthcare worker. 159 responses were eligible for analysis.

The study was deemed exempt from review by the Institutional Review Board (IRB) by the UMass Chan Medical School IRB (Docket #H00018195). As such, the need for informed consent was waived.

### Survey content, data collection, and storage

The first POCT survey was developed in 2019 to assess healthcare professionals’ opinions of POCTs, which specifically compared the responses from providers in cardiovascular medicine to other healthcare professionals. The 2020 survey included additional questions to assess the impact of COVID-19 on healthcare provider impressions of POCT. In 2021, the COVID-19 section was removed and several new questions were added to encapsulate specific issues that arose due to the COVID-19 pandemic. In 2023, additional questions were added to assess the recent rise in artificial intelligence (AI) and machine learning (ML) in POCTs. Due to the omission or addition of specific questions over various iterations, statistical analyses were used only on questions that were identical between the surveys. The complete survey instrument can be found in the S6 Survey.

The survey contained multiple sections, including provider and patient perceptions of POCTs, perceived benefits and concerns, and the most important characteristics of a POCT. Additional closed-ended questions allowed respondents to list up to five conditions that could benefit from POCT, and demographic information including gender, age, race, ethnicity, profession, practice environment, and years in practice. Questions measuring general POCT matters were adapted from the National Heart, Lung, and Blood Institute’s (NHLBI) 2016 Strategic Vision and a survey developed by the researchers from the Point-of-Care Technology Research Network (POCTRN) center at Johns Hopkins University [9, 10]. Questions surrounding business adoption practices of POCTs were adapted from two seminal studies on adopting new technologies [11, 12].

The survey was generated by a Research Electronic Data Capture (REDCap) interface; the secure server is hosted on the UMass Chan network [13]. All data received from participants were transmitted directly into the server for storage and was accessible only by authorized members of the study team.

### Data Analysis

Responses from most questions used a 5-point Likert scale ranging from strongly disagree to strongly agree with the given statement. Survey responses were grouped into two categories for analysis: agreement (responses indicating “Strongly Agree” or “Agree”) and disagreement (responses indicating “Strongly Disagree” or “Disagree”). Checkbox selections for medical specialties were categorized based on an adapted list of standard medical specialties [14].

The importance of various characteristics of POCTs was gathered through a ranking tool. Analysis of important characteristics of POCTs was determined using the following system: 1^st^ most important = 3 points, 2^nd^ most important = 2 points, and 3rd most important = 1 point.

Data analysis was conducted using Excel v2502. Chi-square tests were used to assess differences in responses regarding the benefits and concerns of POCTs. Two-proportion Z-tests were used to identify significant differences in agree proportions between 2019-2021 (aggregate) and 2023. Heat maps were created to visualize the trends in perception of POCTs.

## Results

A total of 159 respondents replied to the 2023 survey. Of those who completed the demographics questions, 49 (48%) identified as male and 51 (46%) as female. Fourteen (13%) respondents were Hispanic or Latino, while 79 (76%) were not. 68 (64%) of the participants self-identified as White, whereas 12 (11%) as Black or African American, 14 (13%) as Asian, 5 (5%) as American Indian or Native Alaskan, and 7 (7%) preferred not to answer. One-quarter of the respondents, 26 (25%), have been in practice for over 20 years, while over half have been in practice for between 0 and 10 years (25 (24%) for 0 – 5 years and 28 (27%) for 6 – 10 years, respectively). Fifty-one (47%) respondents were physicians, 32 (20%) were advanced practice providers, 14 (13%) were registered nurses, and 12 (11%) identified as “other”, including researchers. Three-quarters (79 (76%)) were employed in hospital or ambulatory clinics (Table 1).

**Table 1.**
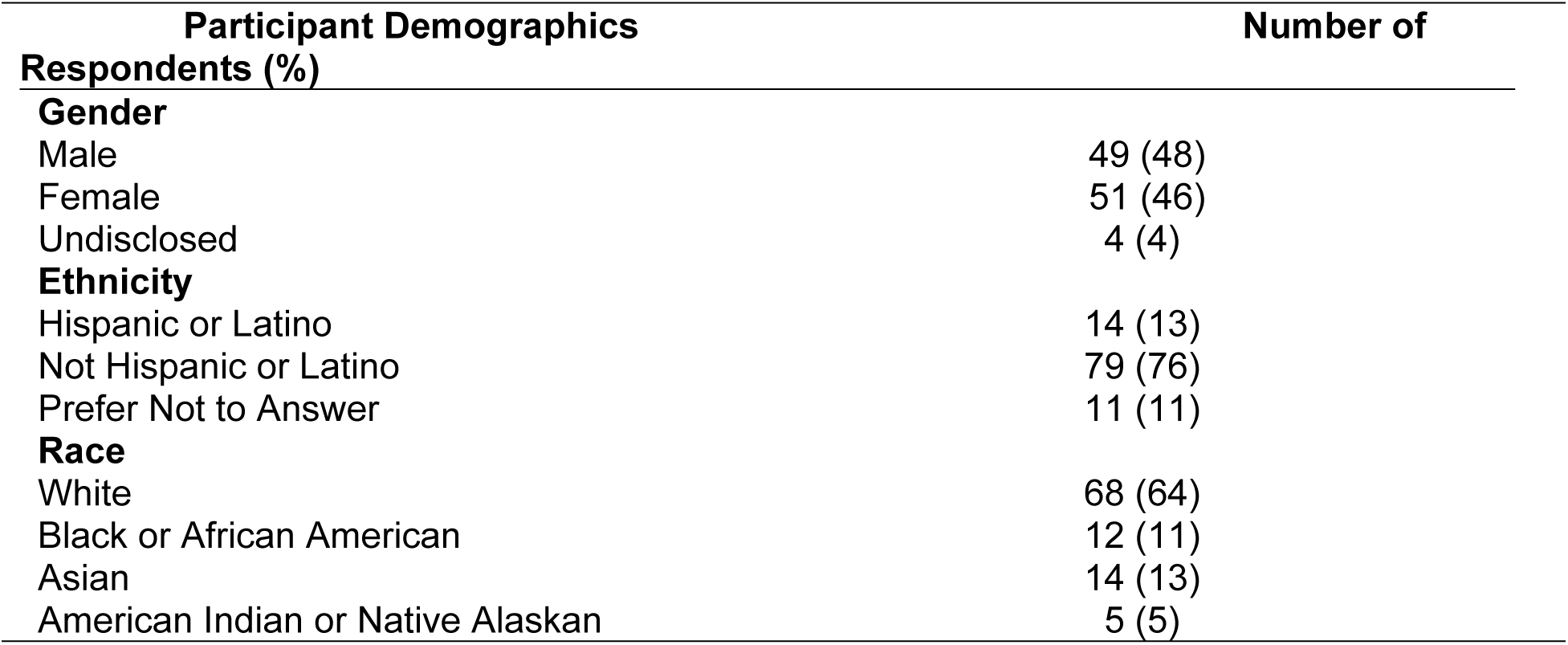
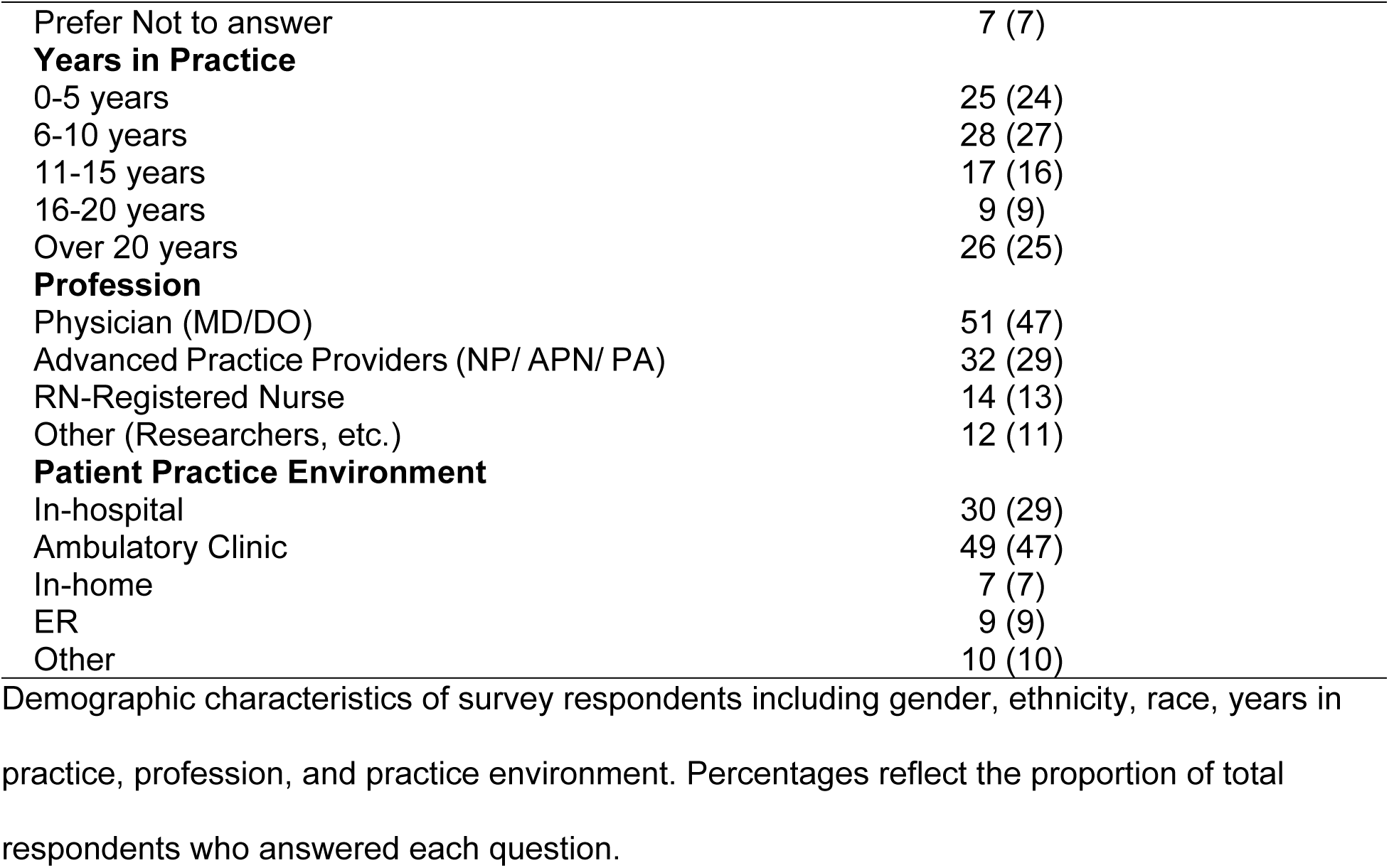
Demographics of Survey Respondents from the 2023 POCT Survey.

Respondents were asked to identify the first, second, and third most important characteristics of POCTs when incorporating them into their current practice (Table 2). In 2023, the three most important characteristics were (1) accuracy, (2) ease of use, and (3) availability. These results are consistent with those of 2020 and 2021, while “does not disturb workflow” was the 3^rd^ most important characteristic in 2019 [1, 3]. The least important characteristics (rank) in 2023 were (11) Device Footprint, (T9) Sample Type, and (T9) CLIA (Clinical Laboratory Improvement Amendments)-Waived Status. Ruggedness was not included in the 2023 survey.

**Table 2.**
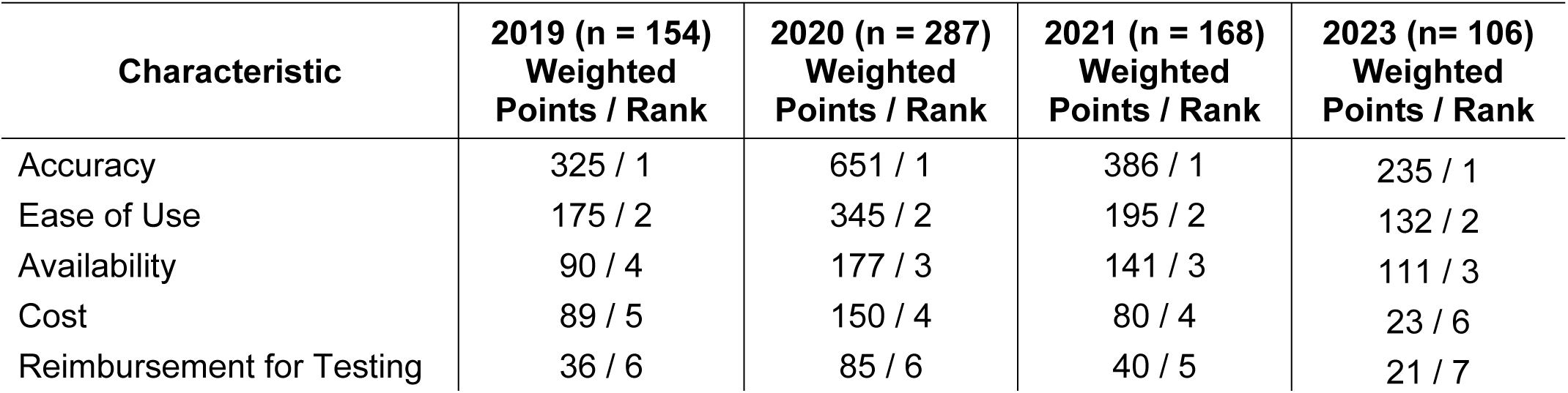
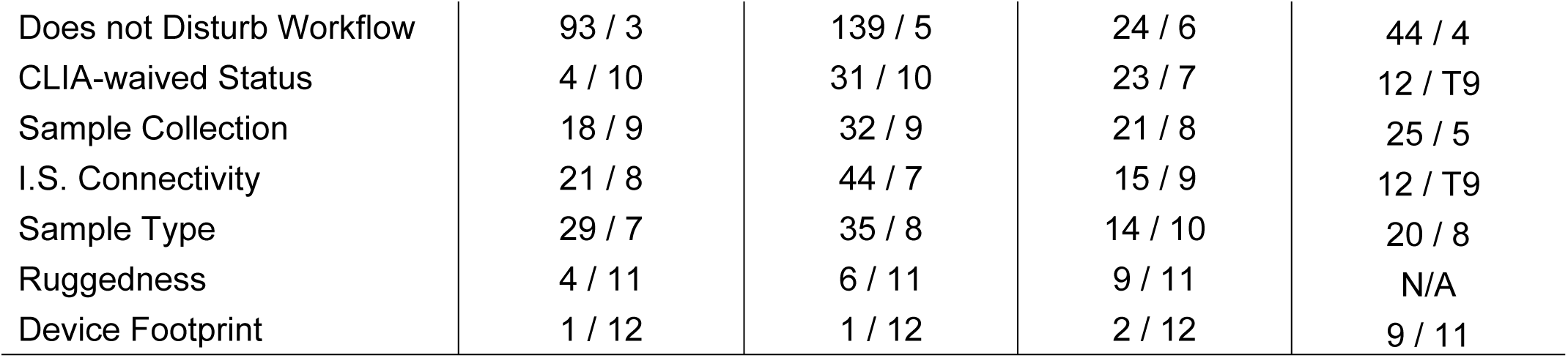
Survey Values* of the Important Characteristics of POCT from Years 2019 – 2023. Ranked importance of POCT characteristics. *Values were determined using a 3-point system (3 points for most important, 2 points for 2^nd^ most important, and 1 point for 3^rd^ most important). Results show comparative longitudinal trends in characteristic prioritization. IS = Information Systems. https://doi.org/10.1371/journal.pone.0299516.t003

### Descriptive Statistics

### 2023 POCTs Benefits and Concerns Comparison to All Prior Years (2019-2021)

The agreement rates on various statements regarding POCTs between the pooled 2019-2021 data and 2023 responses were compared using a two-proportion Z-test (Table 3). Pooled results were used as different response rates were recorded yearly for the survey. Survey respondents were asked to rank their agreement with certain statements: strongly agree, agree, neutral, disagree, or strongly disagree. For this analysis, strongly agree and agree were combined into an agreed category and strongly disagree and disagree were combined into a disagreed category. Cohen’s H effect size was used to measure the magnitude of change, with values above 0.20 indicating small but meaningful differences and a p value of less than or equal to 0.05 indicating statistical significance. The most substantial effect sizes observed were for statements related to confidence in decision making (h = 0.34), and diagnostic certainty (h = 0.31).

**Table 3:**
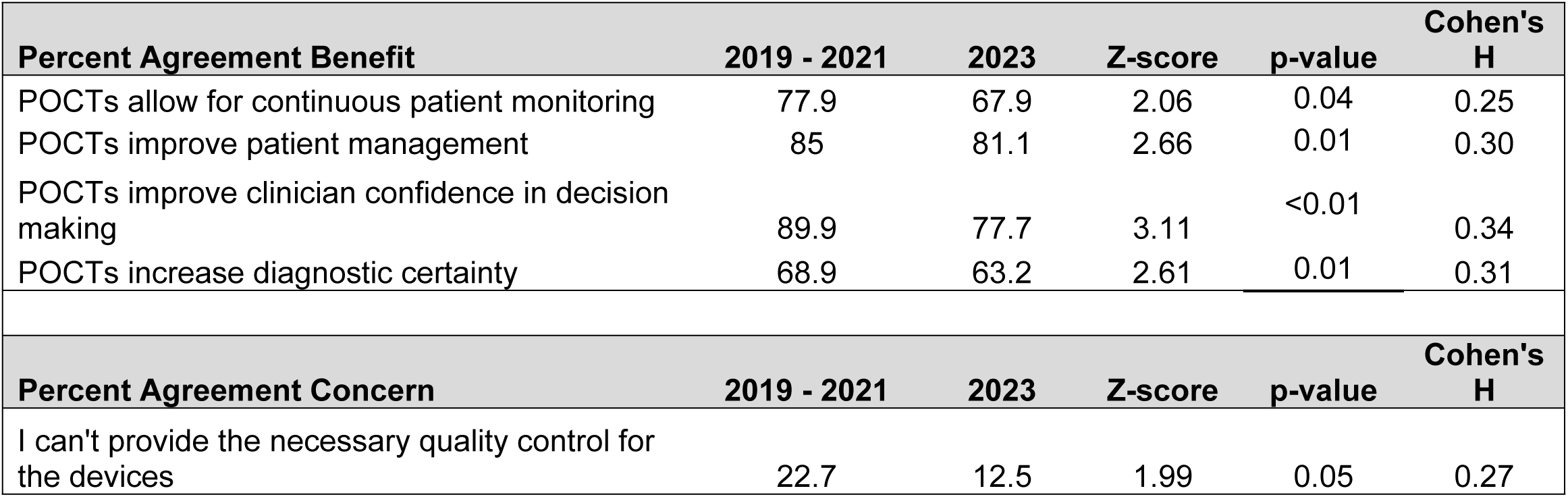
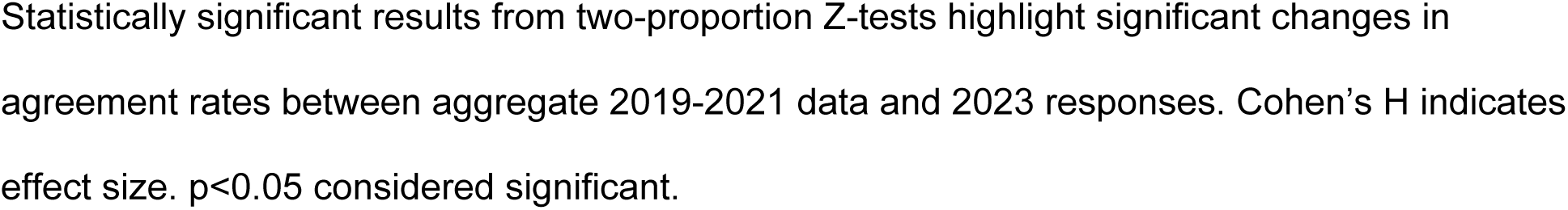
Significant Z-score Results for Benefit and Concern statements 2019-2021 vs. 2023.

For the perceived benefit statements, four of the fifteen statements had a moderate and statistically significant change (h≥0.20 and p <0.05) between the aggregate 2019-2021 and the 2023 responses. Agreement with the statements “POCTs allow for continuous patient monitoring” (Z=2.06, p=0.04, h=0.25), “POCTs improve patient management” (Z=2.65, p=0.01, h=0.30), “POCTs improve clinician confidence in decision making” (Z=3.11, p=<0.01, h=0.34), and “POCTs increase diagnostic certainty” (Z=2.61, p=0.01, h=0.31) significantly decreased in 2023 compared to 2019-2021.

Results from the two-proportion Z-test of POCT concerns differed as there was only one statistically significant increase. The increased agreement with the statement “I can’t provide the necessary quality control for the devices” (Z=1.93, p=0.05, h=0.27) was statistically significant. At the same time, concerns such as “POCTs undermine clinical expertise” (Z=-1.43, p=0.15, h=-0.16) displayed a marginal, non-significant increase. In contrast, the statement “The results of POCTs are difficult to interpret/not definitive” (Z=1.72, p=0.09, h=0.24) showed a marginal, non-significant decrease.

### 2023 POCTs Benefit and Concern Agreement Comparison to 2021

A similar two-proportion Z-test was completed to analyze the agreement rates between the 2023 and 2021 surveys (Table 4). Four of the fifteen showed a statistically significant decrease in perceived benefit statements. Agreement with the statements “POCTs improve patient management” (Z=3.19, p=<0.01, h=0.21), “POCTs improve clinician confidence in decision-making” (Z=3.30, p=<0.01, h=0.22), “POCTs decrease overprescribing of drugs such as antibiotics” (Z=3.44, p=<0.01, h=0.25), and “POCTs increase diagnostic certainty” (Z=2.78, p=<0.01, h=0.20) significantly decreased in 2023 compared to 2021. The most substantial effect sizes observed were for statements related to reimbursement for the cost of the POCT (h = 0.30) and overprescription of drugs (h = 0.25).

**Table 4:** Significant Z-score Results for Benefit and Concern Statements 2021 vs. 2023.

| Percent Agreement Benefit | 2021 | 2023 | Z-score | p-value | Cohen's H |
| --- | --- | --- | --- | --- | --- |
| POCTs improve patient management | 93.8 | 81.1 | 3.19 | $<0.01$ | 0.21 |
| POCTs improve clinician confidence in decision making | 91.9 | 77.8 | 3.30 | $<0.01$ | 0.22 |

| POCTs decrease overprescribing of drugs such as antibiotics | 76.9 | 57.0 | 3.44 | <0.01 | 0.25 |
| --- | --- | --- | --- | --- | --- |
| POCTs increase diagnostic certainty | 78.8 | 63.2 | 2.78 | <0.01 | 0.20 |
| Percent Agreement Concern | 2021.0 | 2023.0 | Z-score | p-value | Cohen's H |
| I might not be reimbursed for the cost of the POCT | 36.7 | 23.1 | 2.33 | 0.02 | 0.30 |

Statistically significant results from two-proportion Z-tests highlight significant changes in agreement levels between 2021 and 2023 responses. Cohen's H indicates effect size. $p < 0.05$ considered significant.

Analysis of the agreement rates regarding perceived concerns yielded one statistically significant decrease in the statement “I might not be reimbursed for the cost of the POCT” (p=0.02, Z=2.33, h=0.30). Notable non-significant decreases included the statements “I can’t provide the necessary quality control for the devices” (Z=1.88, p=0.06, h=0.25) and “The results of POCTs are difficult to interpret/not definitive” (Z=1.90, p=0.06, h=0.25). At the same time, there was a notable non-significant increase for the statement “POCTs undermine clinical expertise” (Z=-1.63, p=0.10, h=-0.20).

#### POCT Expected and Achieved measurements for the 2023 Survey

A Chi-Square test for independence was conducted to examine whether response distributions differed significantly across the years between the categories of agree, neutral, and disagree categories for multiple benefit and concern statements (S1 Table). The highest contributions amongst the benefits statements included diagnostic certainty (x^2^=18.50), patient management (x^2^=9.34), and patient engagement and satisfaction (x^2^=7.52). The highest contributions amongst the concerns statements included the difficulty of interpreting test results (x^2^=26.18), cost concerns (x^2^=20.39), and over-reliance on POCTs (x^2^=16.34). The lone non-significant statement was “POCTs improve patient engagement and buy-in satisfaction” (p=0.08).

#### Heat Map Analysis

Heat maps were developed to visualize the trends in agreement of statements of benefit and concern. Table 5 illustrates the longitudinal responses to the statements of benefit. The color gradient, ranging from white (lowest perceived benefit) to dark blue (highest perceived benefit), demonstrates the year-over-year trends in perceived benefits. The median of each individual statement was set as the mean of the percentage agreement of the aggregate 2019-2021 responses.

**Table 5:**
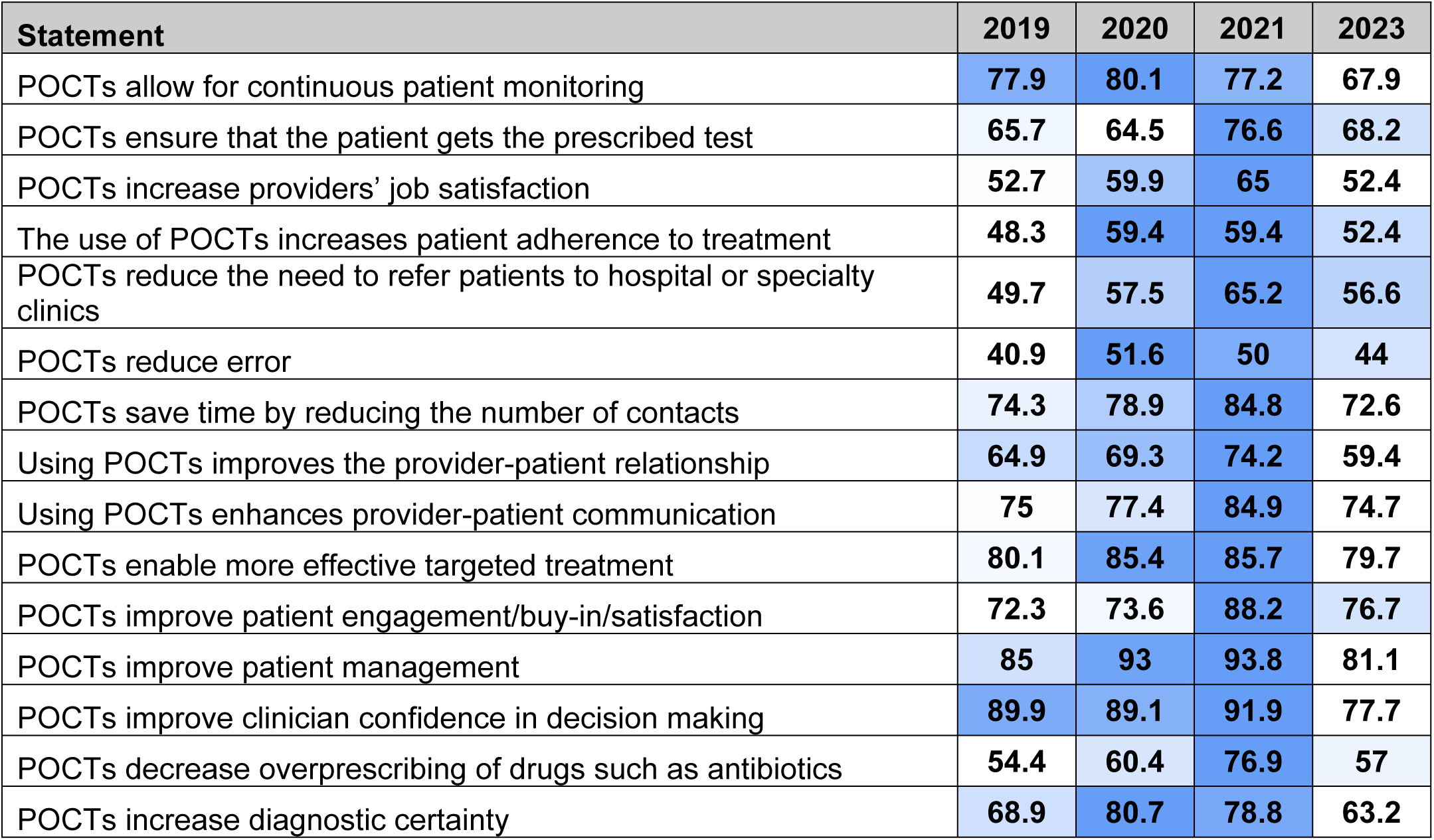
Heat Map of Agreement with Perceived POCT Benefits (2019-2023). Percentage agreement across survey years is visualized with a color gradient from white (lowest percent agreement) to dark blue (highest percent agreement). Results indicate a moderate rise from 2019-2020, peak in 2021, and a decline in 2023.

| Statement | 2019 | 2020 | 2021 | 2023 |
| --- | --- | --- | --- | --- |
| POCTs allow for continuous patient monitoring | 77.9 | 80.1 | 77.2 | 67.9 |
| POCTs ensure that the patient gets the prescribed test | 65.7 | 64.5 | 76.6 | 68.2 |
| POCTs increase providers' job satisfaction | 52.7 | 59.9 | 65 | 52.4 |
| The use of POCTs increases patient adherence to treatment | 48.3 | 59.4 | 59.4 | 52.4 |
| POCTs reduce the need to refer patients to hospital or specialty clinics | 49.7 | 57.5 | 65.2 | 56.6 |
| POCTs reduce error | 40.9 | 51.6 | 50 | 44 |
| POCTs save time by reducing the number of contacts | 74.3 | 78.9 | 84.8 | 72.6 |
| Using POCTs improves the provider-patient relationship | 64.9 | 69.3 | 74.2 | 59.4 |
| Using POCTs enhances provider-patient communication | 75 | 77.4 | 84.9 | 74.7 |
| POCTs enable more effective targeted treatment | 80.1 | 85.4 | 85.7 | 79.7 |
| POCTs improve patient engagement/buy-in/satisfaction | 72.3 | 73.6 | 88.2 | 76.7 |
| POCTs improve patient management | 85 | 93 | 93.8 | 81.1 |
| POCTs improve clinician confidence in decision making | 89.9 | 89.1 | 91.9 | 77.7 |
| POCTs decrease overprescribing of drugs such as antibiotics | 54.4 | 60.4 | 76.9 | 57 |
| POCTs increase diagnostic certainty | 68.9 | 80.7 | 78.8 | 63.2 |

Table 6 illustrates responses to the statements of concern. The color gradient, ranging from white (lowest perceived concern) to dark blue (highest perceived concern), demonstrates the year-over-year trends in perceived concerns. The median of each individual statement was set as the mean of the percentage agreement of the aggregate 2019-2021 responses.

**Table 6:**
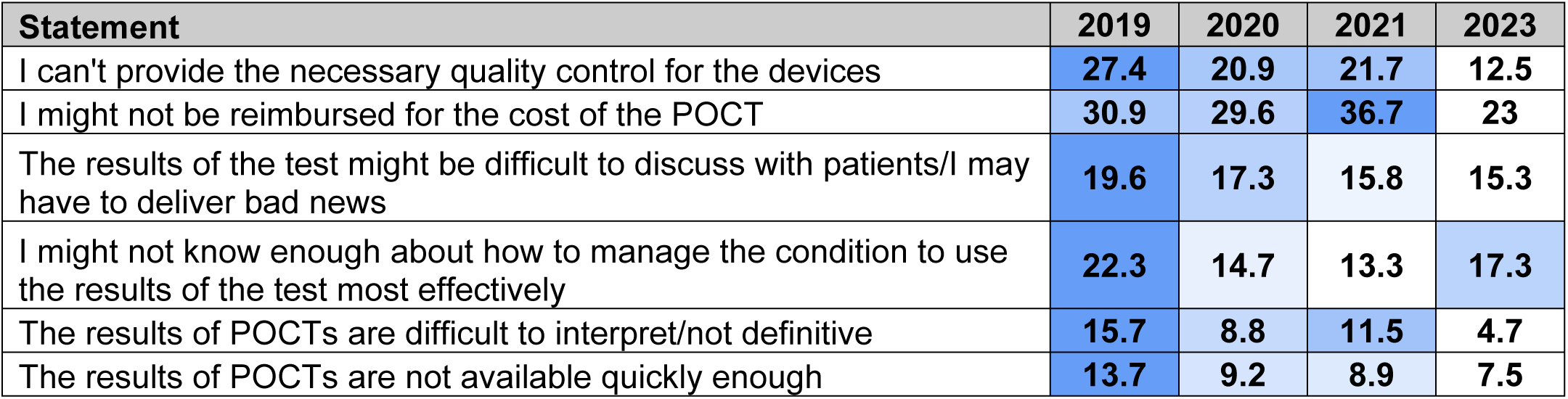
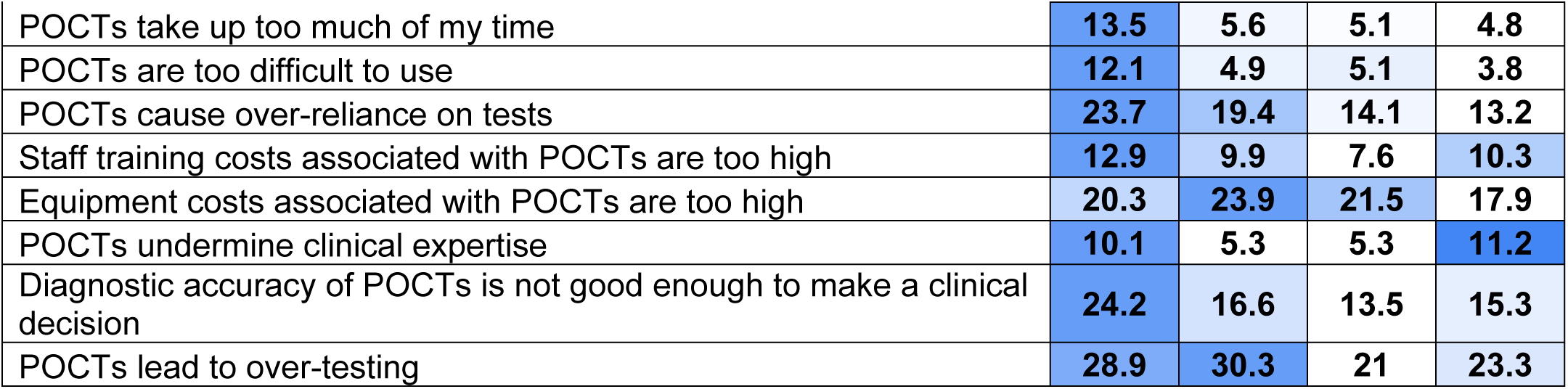
Heat Map of Agreement with Perceived POCT Concerns (2019-2023). Percentage agreement across survey years is visualized with a color gradient from white (lowest percent agreement) to dark blue (highest percent agreement). Results indicate high levels of concern in 2019, a moderate taper through 2021, and the lowest levels of concern in 2023.

#### Pilot questions: Artificial Intelligence and Machine Learning

Three pilot questions surrounding the use of AI and ML were introduced in the 2023 study. Thirty-two percent (n=35) of respondents agreed and 19% (n=20) disagreed with the statement “I am likely to adopt a POCT if it utilizes AI/ML”. Fifty-one percent (n=55) of respondents agreed and 6% (n=7) disagreed with the statement “Use of AI/ML in POCT testing can provide novel information that is not currently available”, while 46% (n=49) agreed and 9% (n=10) disagreed with the statement “My practice is eager to adopt AI/ML learning powered innovations in healthcare.” (Table 7)

**Table 7:**
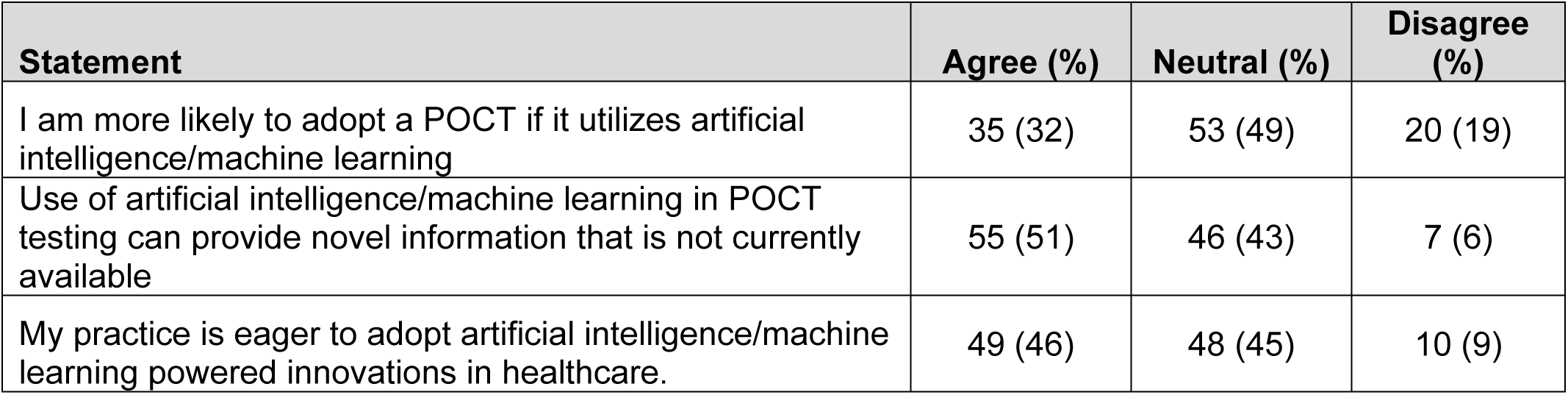
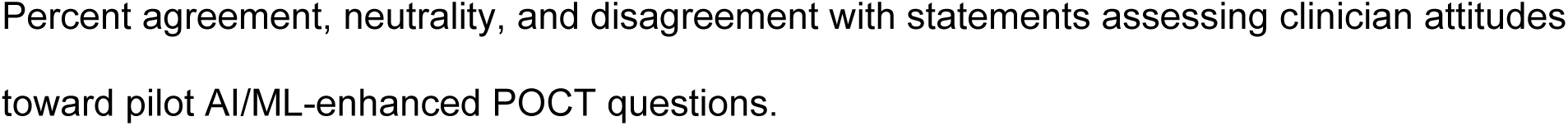
Pilot Artificial Intelligence and Machine Learning Questions.

## Discussion

The 2023 clinician-facing POCT survey highlights significant shifts in healthcare providers’ perceptions and opinions about POCT usage, implementation, and efficacy. The findings demonstrate a tempered perspective, characterized by a reduction in both perceived concerns and perceived benefits. Notably, there was a decline in agreement with several benefit-related statements, indicating reduced confidence in the clinical utility of POCTs. Decreased agreement with statements including “POCTs allow for continuous patient monitoring” and “POCTs improve patient management” may reflect a growing skepticism about the role of POCTs in clinical workflows. This shift in perception could suggest that, with more experience, providers are encountering limitations, such as inconsistent or inaccurate patient use, which may undermine the clinical effectiveness of continuous patient monitoring and management. It may also reflect broader trends in healthcare technology and rising expectations, prompting clinicians to question the real-time reliability and long-term value of these tools.

Perceived benefits peaked in 2021 according to the heat map, with a substantial decrease in the subsequent 2023 survey. Perceived concerns have experienced a more gradual decline throughout the years 2020-2023. One notable statement of concern is “POCTs undermine clinical expertise”, which more than doubled in agreement rate (5.3% in 2020 and 2021 to 11.2% in 2023). In conjunction with the statistically significant decreases in agreement with the statements “POCTs improve clinician confidence in decision-making” and “POCTs decrease overprescribing of drugs such as antibiotics”, the results suggest that POCTs are now viewed with more caution. Possible explanations may include providers growing more critical of POCTs as they continue to integrate and interact with them in routine care workflows, or a heightened awareness of their limitations in clinical settings as their weaknesses become more apparent.

However, it is important to note that not all areas of POCT perception have declined. Healthcare providers continue to value POCTs’ interpersonal aspects, as evidenced by non-significant changes related to improving the provider-patient relationship and enhancing communication. The stability in perceived interpersonal benefits demonstrates the continued relevance of POCTs in enhancing patient engagement and satisfaction, a critical aspect of healthcare delivery.

Concerns about POCTs have evolved. Several concerns including “POCTs take up too much of my time” and “POCTs are difficult to use” showed non-significant changes, while concerns about the POCTs undermining clinical expertise have increased. These shifts in concerns may be indicative of a growing complexity related to integrating POCTs into clinical workflows as they are more widely used, along with the challenge of accurate results and maintaining clinical autonomy when necessary.

The decrease in concerns related to the reimbursement for POCTs may suggest that clinicians are facing fewer financial obstacles to POCT implementation. This contrasts with the high levels of concern regarding reimbursement in the prior 2021 survey [1]. This change could indicate that clinicians are either more familiar with reimbursement policies or that financial barriers to POCT adoption have decreased with time.

While our study highlights important trends surrounding the perceived benefits and concerns of POCTs, some limitations must be considered. The sample size was relatively small for the 2023 survey compared to the aggregate data from previous years, which may impact the robustness of our results. Additionally, 49 participants did not complete the demographic questions, limiting our ability to analyze responses by provider characteristics. In future surveys, these questions should be required while offering a “prefer not to answer” option to respect participant privacy while balancing data completeness. Additional qualitative research may uncover the underlying factors influencing these shifting attitudes. Building off the successful AI/ML pilot questions, our future survey will focus on AI/ML technologies, perceptions, integration practices, and trust. We aim to reach a more diverse healthcare population and expand the settings surveyed to strengthen the generalizability of these findings.

## Conclusion

While POCTs continue to be an asset in clinical settings, the findings of this study suggest a shift in provider attitudes toward a more neutral standpoint – fewer perceived benefits and fewer perceived concerns when compared to three previous surveys between 2019-2021. As the healthcare ecosystem continues to evolve, it will be crucial to address these concerns and explore strategies to bolster confidence in POCTs while keeping patient safety and improved outcomes at the forefront. The three most important characteristics of POCTs have remained consistent throughout the 2020, 2021, and 2023 surveys with accuracy, ease of use, and availability being most highly valued. As POCTs and AI/ML-bolstered technologies continue to infiltrate the healthcare ecosystem and clinical workflows, it is crucial to understand clinician perspectives of such tools continuously. Future surveys should include additional AI and ML-centered questions, acknowledging the recent boom in technological advancements, and targeting more diverse and geographically widespread regions to gain a more comprehensive understanding of clinicians’ viewpoints.

## Data Availability

All relevant data are within the manuscript and its Supporting Information files.

## Acknowledgements

The authors recognize the contributions of Program Officers Jue Chen and Emrim Horgusluoglu from the National Heart, Lung, and Blood Institute and National Center for Complementary and Integrative Health, respectively that contributed to the quality of the survey.

## Author Contributions

**Conceptualization:** Trevor Vigeant, Nathaniel Hafer, Taylor Orwig.

**Data Curation:** Taylor Orwig, Trevor Vigeant, Reilly Riddell.

**Formal Analysis:** Grace White, Trevor Vigeant, Ziyue Wang, Matheus Montenario.

**Funding acquisition:** David D. McManus, Nathaniel Hafer, Bryan Buchholz.

**Investigation:** David D. McManus, Nathaniel Hafer, Bryan Buchholz, Denise Dunlap.

**Methodology:** Taylor Orwig, Bernard Ofosuhene, Nathaniel Hafer, Denise Dunlap.

**Project administration:** Nathaniel Hafer.

**Software:** Ziyue Wang.

**Supervision:** Nathaniel Hafer, Ayorkor Gaba.

**Validation:** Nathaniel Hafer, Ayorkor Gaba.

**Visualization:** Grace White, Trevor Vigeant.

**Writing – original draft:** Trevor Vigeant, Reilly Riddell, Bernard Ofosuhene, Grace White, Matheus Montenario.

**Writing – review & editing:** Nathaniel Hafer, Ayorkor Gaba, Heaven Tatere, Denise Dunlap.

## Supporting Information

**S1 Table**

**S2 Table**

**S3 Table**

**S4 Table**

**S5 Table**

**S6 Survey**

**S7 Survey**

**S8 Survey**

**S9 Survey**

## Notes

### Competing Interest Statement

The authors have declared no competing interest.

### Funding Statement

Yes

### Author Declarations

UMass Chan Medical School IRB

